# Feasibility and Safety in health-related field-based physical fitness tests in adult population: The ADULT-FIT project

**DOI:** 10.1101/2025.07.27.25332266

**Authors:** Carolina Cruz-León, Nuria Marín-Jiménez, Sandra Sánchez-Parente, Milkana Borges Cosic, Alberto Grao-Cruces, José Castro-Piñero, Magdalena Cuenca-García

## Abstract

Studies on feasibility and safety of health-related field-based physical fitness tests in adults are limited, with a lack of methodological homogeneity in the definition of their items. The aim of the present study was to assess the feasibility and safety of the most frequently used health-related field-based physical fitness tests in adult population.

A total of 390 participants, homogeneously distributed by sex, age and physical activity level, were included in the study. Feasibility and safety items proposed from the scientific evidence were assessed for the most frequently used health-related field-based physical fitness tests in adult population. Overall, feasibility items presented a successful answer of 100% in all tests; “participants evaluated” were the 100% of the sample, with an adequate “ratio participants/evaluators” and “time of preparation” (i.e., ∼1 minute). Safety items presented a successful answer of 99% in all tests, “heart rate” (HR) was evaluated only in cardiorespiratory fitness tests; the 97% of the participants reached the “85% HRmax” in the 20-m shuttle run test. For “rate of perceived exertion”, the 51% and 88% of the participants reported a moderate and hard to maximum effort in the 30-s sit-to-stand and the front plank tests, respectively. For “tibial pain” the 32% of the participants reported pain in the 2-km walk test, and for “delayed-onset muscle soreness” the 45% of the participants expressed some degree of muscle soreness, with the 95% of them reporting that cardiorespiratory fitness tests could be the cause.

The most frequently used health-related field-based physical fitness tests are feasible and safe in adult population homogeneously distributed by sex, age and physical activity level.

## Introduction

Health-related physical fitness has been shown to have great benefits in prevention and prognosis of several age-related diseases or pathologies in the adult population [1–3]. The health-related physical fitness components are cardiorespiratory fitness, musculoskeletal fitness (i.e., muscular fitness and flexibility), motor fitness and body composition [4]. Cardiorespiratory fitness has been associated with the reduction of the risk to develop of non-communicable diseases like obesity [5], diabetes [6] or metabolic syndrome [7]. Muscular fitness is related to the reduction of obesity [8], cardiovascular and metabolic diseases [9], improvement of bone health [10, 11] and prevents from all-cause mortality [3]. Appropriate motor fitness level is associated with a greater decrease in disability, reduced risk of falls and frailty, depression [12], cognitive decline [13] and cardiovascular disease and all-cause mortality [2, 14]. Finally, body composition is inversely related to numerous health problems in youth and adulthood [15].

Therefore, physical fitness assessment is a key method of health prevention and diagnosis. In this sense, health-related physical fitness can be objectively assessed by laboratory devices (i.e., reference methods or gold standards) in a safe and controlled environment. Regardless, its use is limited due to qualified technicians, specific instruments and facilities required. On the other hand, health-related field-based physical fitness tests are shown to be time-efficient, low-cost, and an easy to administer option that allow to measure a large number of people at the same time [16].

A health-related field-based physical fitness test battery must address a number of fundamental requirements to be used: i) predictive validity, ii) criterion-related validity, iii) reliability, iv) feasibility, v) safety, and vi) responsiveness whether in clinical settings, sport centers, schools and any possible environment in epidemiological studies [17].

There is available evidence on predictive validity [1–3], criterion-related validity [18], and reliability [19] of field-based physical fitness tests in adults. However, studies on feasibility and safety of health-related field-based physical fitness tests in adults are limited, with a lack of methodological homogeneity in the definition of the feasibility and safety items [20]. Feasibility is the ability of a test to function properly outside the controlled environment of a laboratory and must comply with various elements such as test facility, time required, equipment, evaluators, among others. Feasibility allows researchers to evaluate whether ideas and outcomes can be adapted to be relevant and sustainable [21]. Otherwise, safety refers to the assessment of risk or harm that is likely to occur before, during and after the performance of the tests. A test is considered safe when it does not cause physical complications to participants [22].

Given that it is known which health-related field-based physical fitness tests are valid and reliable in adults, it would be advisable to also know which health-related field-based physical fitness are feasible and safe in this population, to be able to propose, a health-related field-based physical fitness test battery that accomplish with the aforementioned precepts. Therefore, the aim of the present study was to assess the feasibility and safety of the most frequently used health-related field-based physical fitness tests in adult population.

## Methods

### Study sample and design

The present study is part of a national project: the ADULT-FIT study (DEP2017-88043- R), whose principal objective was to propose a health-related field-based physical fitness tests battery in adult population, based on predictive and criterion-related validity, reliability, feasibility, safety and responsiveness.

Sample size was determined according to the sampling formula for infinite populations with a two-sided significance level of 5% and a power of 95%. A 10% drop-out rate was considered (G*power 3.1.9.6, Düsseldorf, Germany). Participants were recruited through leaflet, local newspapers and social media from Cadiz. A total of 390 participants were included in the study. The total sample was homogeneously distributed by sex (50.5% women), age (18–34, 35–49, and 50–64 years) and physical activity level (non-active and active).

The inclusion criteria of this study were: i) adults (i.e., 18 to 64 years old), ii) not suffering from any physical or mental illness that impairs their ability to participate in physical activity, iii) willingness to perform all the tests comprising the study, and iv) be able to read and understand the informed consent form and the purpose of the study. The exclusion criteria were: i) severe or end-stage illness, ii) myocardial infarction three months prior to study beginning, iii) unsteady cardiovascular condition, iv) medical indication which prevents the tests from being performed, and v) any injury or circumstance that makes it incompatible with the correct performance of the tests.

All participants provided voluntarily written informed consent to be part of the present study. The study was approved by the Committee for Research of Cadiz, Spain.

After providing written informed consent and being informed of the protocol to be carried out, they signed the “Physical Activity Readiness Questionnaire” [23]. Participants were initially classified as active/non-active according to the World Health Organization’s recommendations for adults [24], once they answered the question: *how many days (in a typical week) do you practice physical activity/exercise or some sport, of at least moderate intensity, lasting at least 60 minutes per day?*

### Health-related field-based physical fitness tests assessment

The health-related physical fitness components measured and the tests related to them were: body composition (i.e., weight, height, triceps and subscapular skinfolds, and neck, waist and hip circumferences), cardiorespiratory fitness (i.e., the 6-min walk, 2-km walk, and 20-m shuttle run tests), muscular fitness (i.e., the handgrip strength, standing long jump, 30-s sit-to-stand, and front plank tests), and motor fitness (i.e., the 6-m gait speed, 2.45-m time up & go, 4x10-m shuttle run, and single-leg stand tests).

Prior to the field-based physical fitness testing sessions, all participants underwent a standardized 10-minute warm-up. All participants received detailed instructions for the tests and were actively encouraged to do their best in each test. In addition, participants were instructed to rest for 24 hours before the assessments, to maintain their nutritional and hydration habits and wear appropriate sport clothes (i.e., sports clothing and sports shoes that allow correct movement and sliding).

The tests were carried out in three non-consecutive days, with at least 72 hours apart, in an indoor/outdoor facility under appropriate conditions (i.e., free space with good lighting and ventilation; no objects restricting the performance of the tests; flat, clean, undamaged, non-slip and non-abrasive surface; and specific test equipment in perfect and non-injurious condition). The tests were assessed and supervised by researchers with experience in field-based physical fitness testing.

The procedure/protocol of field-based physical fitness tests can be consulted in *Supplementary Material 1*.

### Feasibility assessment

The feasibility items proposed, based on scientific evidence [20], were: i) “appropriate facilities” (i.e., the test facility is in optimal conditions in terms of lighting and ventilation and enough space), ii) “appropriate equipment” (i.e., the audio instruments and speakers for the assessment of the tests are working properly), iii) “appropriate sports clothes” (i.e., correct sport clothes and shoes of the participants), iv) “test’s instruction understood” (i.e., the participant understand the test’s instructions), v) “easy to administer” (i.e., participants perform the test without difficulty and correctly, related to: technical performance, modifications to the original protocol or familiarisation session with technical performance; and evaluator-administered tests, recalled data without difficulty and without the need for profound learning about the items to be assessed), vi) “participants evaluated/not evaluated” (i.e., participants who perform the test and participants who did not perform), vii) “ratio participants/evaluators” (i.e., number of evaluators per test to evaluate a certain number of participants), viii) “time of preparation” (i.e., required time to prepare the test), and ix) “time of performance” (i.e., required time for the participant to complete de test).

### Safety assessment

The safety items proposed, based on scientific evidence [20], were: i) “instrument allergy” (i.e., manual dynamometer); ii) “adverse events” (i.e., falls, injury and/or sick feeling) that may occur during the performance of the test; iii) “heart rate” (HR) (registered at the end of the 6-min walk, 2-km walk, and 20-m shuttle run tests); iv) “rate of perceived exertion” (RPE) (assessed in the 30-s sit-to-stand and front plank tests) using the Borg scale of 1 to 10 for self-perception of general exertion [25], where 1 means “very light”, 2-3 “light”, 4-6 “moderate”, 7-8 “hard”, 9 “very hard” and 10 “maximum effort”; v) “tibial pain” (registered at the end of the 2-km walk test) in a scale of 0 to 10 for subjective self-perception [26] (0 means “no pain”, 1-3 “low”, 4-6 “moderate” and 7-10 “severe” tibial pain); and vi) “delayed-onset muscle soreness” (DOMS) [27], through a self-reported questionnaire 48 hours post assessment of the field-based physical fitness tests. The items recorded in the DOMS questionnaire were: a) “did you experienced DOMS posterior to assessment of the field-based fitness test?” (yes/no answers); b) if the answer was yes, “how much pain do you feel?” (options were: “very low”, “low”, “moderate”, “much” and “severe”); c) “where you had the muscle soreness?” (options were: “left arm”, “right arm”, “both arms”, “lower zone back”, “upper zone back”, “lower and upper zones back”, “left leg”, “right leg”, “both legs”, “both arms and legs”, “lower zone back and both legs”, “upper zone back and both legs” and “other”); d) “what test do you think caused your muscle soreness?” (options were: “the muscular fitness tests” (i.e., the handgrip strength, standing long jump, 30-s sit-to-stand and front plank tests) or “the cardiorespiratory fitness tests” (i.e., the 6-min walk, 2-km walk, and 20-m shuttle run tests)); e) “you have difficulty performing activities of daily living due to muscle soreness?” (options were: “none”, “when picking up a heavy object at the level of your abdomen/trunk”, “when picking up objects from a piece of furniture above my head”, “when descending stairs/climbing stairs”, “when bending down to pick up an object” and “other”); and f) “if in the previous question you have indicated other, please specify which on” (open answer).

### Statistical Analysis

An acceptable level of feasibility occurs when the proposed items (i.e., “appropriate facilities”, “appropriate equipment”, “appropriate sport cloths”, “test’s instruction understood”, and “easy to administer”) shows ≥95% positively answers from participants [27], presented as frequency and percentage. The items “participants evaluated/not evaluated” and “ratio participant/evaluator” are expressed as frequency. The “time of preparation” is presented as mean and standard deviation (SD; seconds) and the “time of performance” as minimum and maximum time ranges (minutes or seconds) for each field-based physical fitness test, with the exception of those tests whose time of performance is established by protocol.

An acceptable level of safety occurs when the proposed items (i.e., “instrument allergy” and “adverse events”) shows ≥99% positively answers from participants [27], presented as frequency and percentage. The “HR” is presented in mean (SD) of beats per minute (bpm). Moreover, the “85% HRmax” is presented as the frequency and percentage of participants who reached or exceed their 85% age-predicted maximum HR. The maximum HR was calculated according to Tanaka et al., [28] (i.e., 208 - 0.7 x age). The “RPE”, “tibial pain” and “DOMS” items are presented as frequency and percentage. All the statistical procedures were conducted using SPSS v. 25.0 software for Windows.

### Results Feasibility

#### Body composition

The data collected from the feasibility items for the assessment of body composition are presented in **Table 1**. All the items assessed (i.e., “appropriate facilities”, “appropriate equipment”, “appropriate sport clothes”, “test’s instruction understood”, and “easy to administer”) presented a successful answer of 100%. The “ratio participant/evaluator” was 1/1 for the assessment.

**Table 1.**
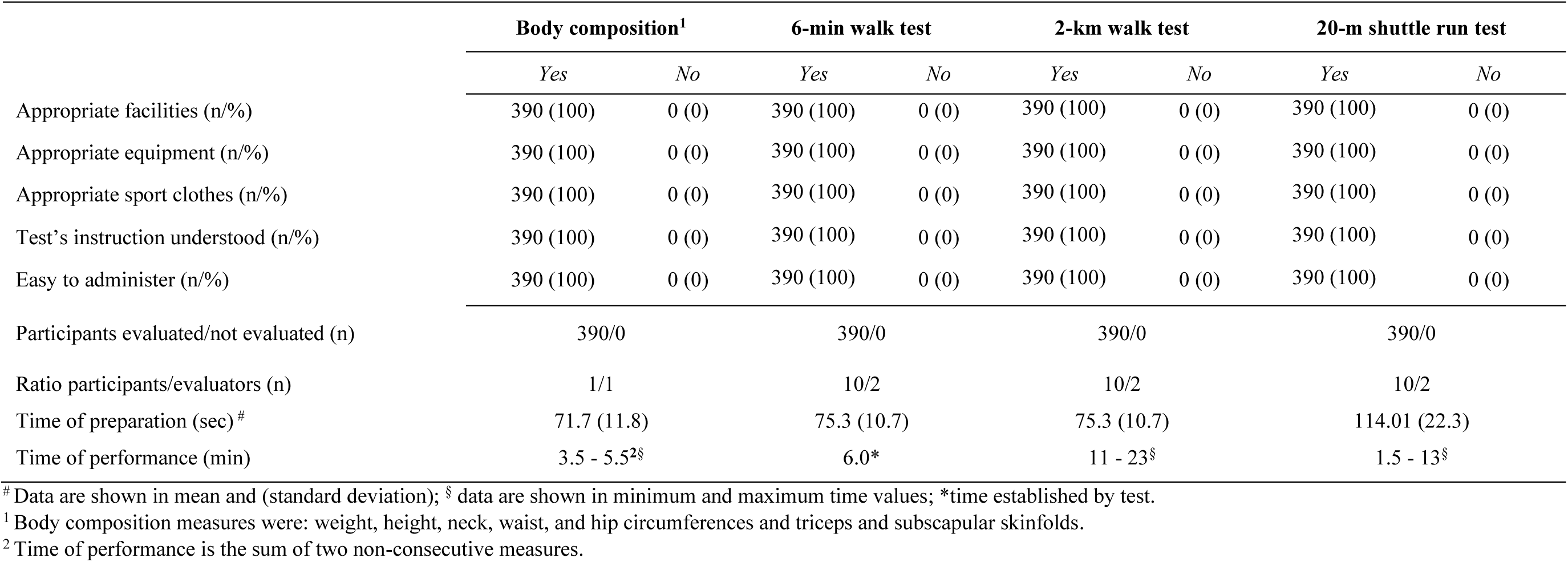
Feasibility of the measurement protocol of field-based body composition and cardiorespiratory fitness tests.

The “time of preparation” of the tests took 71.7 (11.8) seconds and the “time of performance” ranged from 3.5 to 5.5 minutes by participant.

#### Cardiorespiratory fitness tests

The data collected from the feasibility items for the assessment of cardiorespiratory fitness tests are presented in **Table 1**. All the items assessed presented a successful answer of 100%.

The “ratio participant/evaluator” was 10/2 for the correct assessment of the cardiorespiratory fitness tests (i.e., 6-min walk, 2-km walk, and 20-m shuttle run tests). Referring to the item “time of preparation”, the 6-min walk test 75.3 (10.7) seconds, the 2-km walk test took 75.3 (10.7) seconds, and the 20-m shuttle run test 114.01 (22.3) seconds.

The “time of performance” for the 6-min walk test the time was set by the test criterion (i.e., 6 minutes), the 2-km walk test ranged from 11 to 23 minutes. Finally, the time to perform the 20-m shuttle run test ranged from 1.5 to 13 minutes (which also correspond to number of stages).

#### Muscular fitness

The data collected from the feasibility items for the assessment of muscular fitness tests are presented in **Table 2**. All the items assessed presented a successful answer of 100%. The “ratio participant/evaluator” for the handgrip strength and the standing long jump tests was 1/1. Regarding the 30-s sit-to-stand and front plank tests the ratio was 1/2.

**Table 2.** Feasibility of the measurement protocol of field-based muscular and motor fitness tests.

|  | **Handgrip strength** |  | **Standing long jump** |  | **30-s sit-to-stand** |  | **Front plank** |  | **6-m gait speed** |  | **2.45-m time up & go** |  | **4x10-m shuttle run** |  | **Single-leg stand** |  |
| --- | --- | --- | --- | --- | --- | --- | --- | --- | --- | --- | --- | --- | --- | --- | --- | --- |
|  | *Yes* | *No* | *Yes* | *Yes* | *Yes* | *No* | *Yes* | *No* | *Yes* | *No* | *Yes* | *No* | *Yes* | *No* | *Yes* | *No* |
| Appropriate facilities (n/%) | 390 (100) | 0 (0) | 390 (100) | 0 (0) | 390 (100) | 0 (0) | 390 (100) | 0 (0) | 390 (100) | 0 (0) | 390 (100) | 0 (0) | 390 (100) | 0 (0) | 390 (100) | 0 (0) |
| Appropriate equipment (n/%) | 390 (100) | 0 (0) | 390 (100) | 0 (0) | 390 (100) | 0 (0) | 390 (100) | 0 (0) | 390 (100) | 0 (0) | 390 (100) | 0 (0) | 390 (100) | 0 (0) | 390 (100) | 0 (0) |
| Appropriate sport clothes (n/%) | 390 (100) | 0 (0) | 390 (100) | 0 (0) | 390 (100) | 0 (0) | 390 (100) | 0 (0) | 390 (100) | 0 (0) | 390 (100) | 0 (0) | 390 (100) | 0 (0) | 390 (100) | 0 (0) |
| Test's instruction understood (n/%) | 390 (100) | 0 (0) | 390 (100) | 0 (0) | 390 (100) | 0 (0) | 390 (100) | 0 (0) | 390 (100) | 0 (0) | 390 (100) | 0 (0) | 390 (100) | 0 (0) | 390 (100) | 0 (0) |
| Easy to administer (n/%) | 390 (100) | 0 (0) | 390 (100) | 0 (0) | 390 (100) | 0 (0) | 390 (100) | 0 (0) | 390 (100) | 0 (0) | 390 (100) | 0 (0) | 390 (100) | 0 (0) | 390 (100) | 0 (0) |
| Participants evaluated/not evaluated (n) | 390/0 |  | 390/0 |  | 390/0 |  | 390/0 |  | 390/0 |  | 390/0 |  | 390/0 |  | 390/0 |  |
| Ratio participants/evaluators (n) | 1/1 |  | 1/1 |  | 1/2 |  | 1/2 |  | 1/2 |  | 1/2 |  | 1/1 |  | 1/2 |  |
| Time of preparation (s) <sup>#</sup> | 39.1 (9.3) |  | 69.1 (10.3) |  | 60.8 (9.4) |  | 77 (11.7) |  | 55.8 (9.9) |  | 52.3 (8.5) |  | 91.8 (10.3) |  | 55.3 (9.4) |  |
| Time of performance (min or <u>sec</u>) | 1.8 - 3.0<sup>1</sup> §* |  | 1.3 - 2.3<sup>1</sup> §* |  | <u>30</u>** |  | 1.1 - 9.3 §* |  | <u>1.8 - 3.6</u> § |  | <u>4 - 7</u> § |  | 1.2 - 1.3<sup>1</sup> §* |  | 3.0 - 3.6<sup>1</sup> §* |  |

<sup>#</sup> Data are shown in mean and (standard deviation); § data are shown in minimum and maximum time values; \*average minutes per participant; \*\*time established by test.
<sup>1</sup> Time of performance is the sum of two attempts and 1-minute rest between attempts.

The “time of preparation” was 39.1 (9.3) seconds for the handgrip strength test, 69.1 (10.3) seconds for the standing long jump test, 60.8 (9.4) seconds for the 30-s sit-to-stand test, and 77 (11.7) seconds for the front plank test.

Referring to “time of performance” item, the handgrip strength test ranged from 1.8 to 3 minutes (including two attempts and 1-minute rest between attempts), the standing long jump test, from 1.3 to 2.3 minutes (also including two attempts and 1-minute rest between attempts) and the front plank test, from 1.1 to 9.3 minutes. In the 30-s sit-to-stand test, the time to perform the test was set by the test criterion (i.e., 30 seconds).

#### Motor fitness

The data collected from the feasibility items for the assessment of motor fitness tests are presented in **Table 2**. All the items assessed presented a successful answer of 100%. For the proper assessment of the tests, the “ratio participant/evaluator” was 1/2 for the 6-m gait speed, 2.45-m time up & go and single-leg stand tests, and 1/1 for the 4x10-m shuttle run test.

Regarding “time of preparation” for the 6-m gait speed test was 55.8 (9.9) seconds, 52.3 (8.5) seconds for the 2.45-m time up & go test, 91.8 (10.3) seconds for the 4x10-m shuttle run test, and 55.3 (9.4) seconds for single-leg stand test.

Regarding to the “time of performance” item, the 6-m gait speed test ranged from 1.8 to 3.6 seconds, the 2.45-m time up & go test ranged from 4 to 7 seconds, the 4x10-m shuttle run test ranged from 1.2 to 1.3 minutes, and for the single-leg stand test the time was already stablished by the test criterion; however, the assessment took between 3 to 3.6 minutes per participant (including 30 seconds for each leg and 1-minute rest between eyes open and eyes closed modality).

## Safety

The data collected from the safety items for the assessment of the health-related field-based physical fitness tests answered are presented in **Table 3**. No complications occurred during the testing procedure referred to “instruments allergy” and/or “adverse events” (i.e., falls, injury or sick feeling), except one participant (0.3%) that felt sick at the end of the 20-m shuttle run test.

**Table 3.** Safety of the measurement protocol of field-based fitness tests answered by evaluators and participants.

| Field-based physical fitness test | Instrument allergy | Adverse events |  |  | HR<sup>§</sup> | 85% HRmax | RPE<sup>§</sup> | Tibial pain<sup>#</sup> | DOMS* |
| --- | --- | --- | --- | --- | --- | --- | --- | --- | --- |
|  | n (%) | Falls<br>n (%) | Injury<br>n (%) | Sick feeling<br>n (%) | Mean (SD) | n (%) | n (%) | n (%) | n (%) |
| Body composition | 0 (0) | 0 (0) | 0 (0) | 0 (0) | n/a | n/a | n/a | n/a | n/a |
| 6-min walk | 0 (0) | 0 (0) | 0 (0) | 0 (0) | 134.9 (9.3) | 28 (7.2) | n/a | n/a |  |
| 2-km walk | 0 (0) | 0 (0) | 0 (0) | 0 (0) | 144 (7.1) | 125 (32.1) | n/a | No pain: 266 (68.2)<br>Pain: 124 (31.8): low: 46 (11.8), moderate: 43 (11), severe: 35 (9) | 162 (95.3)<sup>1</sup> |
| 20-m shuttle run | 0 (0) | 0 (0) | 0 (0) | 1 (0.3) | 169.2 (5.7) | 378 (97.0) | n/a | n/a |  |
| Handgrip strength | 0 (0) | 0 (0) | 0 (0) | 0 (0) | n/a | n/a | n/a | n/a | n/a |
| Standing long jump | 0 (0) | 0 (0) | 0 (0) | 0 (0) | n/a | n/a | n/a | n/a | 8 (4.7) |
| 30-s sit-to-stand | 0 (0) | 0 (0) | 0 (0) | 0 (0) | n/a | n/a | Very light/light: 153 (39.2)<br>Moderate: 200 (51.3)<br>Hard: 37 (9.5) | n/a | n/a |
| Front plank | 0 (0) | 0 (0) | 0 (0) | 0 (0) | n/a | n/a | Moderate: 46 (11.8)<br>Hard: 237 (60.8)<br>Very hard: 97 (24.9)<br>Maximum effort: 10 (2.5) | n/a | n/a |
| 6-m gait speed | 0 (0) | 0 (0) | 0 (0) | 0 (0) | n/a | n/a | n/a | n/a | n/a |
| 2.45-m time up & go | 0 (0) | 0 (0) | 0 (0) | 0 (0) | n/a | n/a | n/a | n/a | n/a |
| 4x10 shuttle run | 0 (0) | 0 (0) | 0 (0) | 0 (0) | n/a | n/a | n/a | n/a | n/a |
| Single-leg stand | 0 (0) | 0 (0) | 0 (0) | 0 (0) | n/a | n/a | n/a | n/a | n/a |

DOMS: Delayed-onset muscle soreness; HR: heart rate; HR max.: maximum heart rate; RPE: Rate of perceived exertion; SD: standard deviation.
<sup>§</sup> RPE Borg scale (range 1-10): very light/light (1-3), moderate (4-6), hard (7-8), very hard (9), and maximum effort (10).
<sup>#</sup> Tibial pain self-perception (scale range 0-10): no pain (0), low (1-3), moderate (4-6), and severe (7-10) tibial pain.
\* DOMS self-reported questionnaire answered by participants 48 hours post evaluation of the field-based physical fitness tests.
<sup>1</sup> Participants reported that cardiorespiratory fitness tests (i.e., the 6-min walk, the 2-km walk, and the 20-m shuttle run tests) could be the cause of their DOMS.

The mean (SD) value of “HR” in the 6-min walk test was 134.9 (9.3) bpm, 144 (7.1) bpm for 2-km walk test, and 169.2 (5.7) bpm for 20-m shuttle run test. None of the participants reached its maximum HR in the 6-min walk and 2-km walk tests; however, the 22.8% of participants reached their maximum HR in the 20-m shuttle run test.

Regarding the “85%HRmax” item, only the 7% of the participants reached it in the 6-min walk test. From this percentage, the 79% were females, the 32% were physically active and belonged to two age groups (7% for 35-49 years and 93% for 50-64 years). For the 2-km walk test, the 32% of the participants reached their “85%HRmax”, from this percentage the 59% were females, the 45% were physically active and belonged to two age groups (26% for 35-49 years and 74% for 50-64 years). For the 20-m shuttle run test, the 97% of the participants reached their “85%HRmax”, this percentage was balanced by sex (50%) and physical activity level (50%), but not by age groups (28% for 18-34 years, 36% for 35-49 years and 36% for 50-64 years).

For “RPE” item, the 39.2% of participants reported a “very light/light”, 51.3% a “moderate” and 9.5% a “hard” effort in the 30-s sit-to-stand test. The 11.8% of participants reported a “moderate”, 60.8% a “hard”, 24.9% a “very hard”, and 2.5% a “maximum effort” in the front plank test.

The 31.8% of the participants expressed having felt pain associated with the anterior tibial muscles at the end of the 2-km walk test. Of them, the 37.2% reported “low”, 34.6% “moderate”, and 28.2% “severe” tibial pain.

The “DOMS” questionnaire had a response rate of 381 participants (97.7%). The 44.6% of the participants expressed some degree of “DOMS”. Among the responders, the 58.8% experienced “very low/low”, 25.3% “moderate”, 12.9% “quite”, and the 2.9% “severe” degree. Moreover, the 89.4% of the sample that answered the questionnaire, reported pain in “both legs”, 6.5% in the “lower zone back and both legs”, and the 4.1% reported pain in “both arms and legs”. The 95.3% of participants reported that cardiorespiratory fitness tests (i.e., the 6-min, the 2-km walk, and the 20-m shuttle run tests) could be the cause of their “DOMS”; however, the 4.7% of participants reported that the standing long jump test could be the cause. And finally, regarding to the difficulty, they may have experienced in carrying out daily activities due to muscular soreness, the 18% reported difficulties associated to this. Of those who did experience difficulties, the 58% reported that felt them when they “descending stairs/climbing stairs”, the 28% reported “when bending down to pick up an object”, and 14% reported “other” (i.e., difficult to walk normally).

## Discussion

The main findings of the present study suggest that the most frequently used health-related field-based physical fitness tests are feasible and safe in adult population homogeneously distributed by sex, age, and physical activity level.

### Feasibility

All participants completed the proposed field-based physical fitness tests without any difficulties. All the proposed items presented a successful answer of 100% by the participants. The items “appropriate facilities”, “appropriate equipment” and “appropriate sports clothes” should be implicitly included in the assessment of health-related field-based physical fitness tests; however, we have included them based on a previous study in children and adolescents [27], which results are in line with our findings.

Regarding to the items “test’s instruction understood” and “easy to administer”, several studies presented a similar successful answer in these items in children and adolescents [29–33] and adults [22]. However, one study in children and adolescents [27], where the assessments were conducted by Physical Education teachers, observed difficulties in the evaluation of the body composition, specifically with the skinfold measurements due to unfamiliarity. On the contrary, in our study, the evaluators had previous experience. In another study in preschoolers [34], difficulties in the standing long jump were observed, especially in the youngest (3-year-old group). This might have occurred because children do not have a developed jumping pattern, and in our case, we approached the standing long jump as a natural movement and not as a specific skill. Regarding the “participants evaluated/not evaluated” item, in our study all participants performed the tests. Other studies also had a high performance rate, between 95-100% in children and adolescents [30–32, 35], adults [22, 36, 37], and older participants [38]. Concerning to the “ratio participants/evaluators” item, in the cardiorespiratory fitness tests was 10 participants by 2 evaluators. Previous studies in preschoolers showed an optimal ratio of 4 to 8 participants by 2 evaluators (1 of them running with the children to set the pace) to assess the 20-m shuttle run test [33, 34]; while in children and adolescents an optimal ratio was 20 participants by 1 evaluator [27]. The “ratio participants/evaluators” most used in muscular and motor fitness tests were 1/1 and 1/2, respectively. Other studies also showed 1-2 evaluator to assess the handgrip strength [27, 34], the standing long jump [27, 34] and the 4x10-m shuttle run tests [34] in children and adolescents. Only one study [22] mentioned that 1-3 evaluators were needed for the assessment of the muscular and motor fitness tests in adults. We believe that it is important to include 2 evaluators in the muscular fitness and motor fitness tests in order to have a correct measurement (i.e., registration of data, better control of correct execution of those tests with technical complexity and more safety), especially when assessing adult population with low physical fitness.

The “time of preparation” was approximately 1 minute for the majority of the tests except, for the handgrip strength test, which showed the shortest time, which corresponded mainly to the time needed to measure the hand size in women. The 20-m shuttle run test took ∼2 minutes of preparation, in concordance with Cadenas-Sánchez et al., [33] which took ∼3 minutes in preschoolers, although España-Romero et al., [27] took ∼5 minutes in children and adolescents. We could consider that all the tests were easy to prepare and the time that takes to prepare them is adequate.

For the “time of performance” item, we took between 3.5 to 5.5 minutes per participant in body composition assessments (i.e., weight, height, neck, waist, and hip circumferences and triceps and subscapular skinfolds); similar to the study by España-Romero et al., [27] in which 5 minutes were required per child or adolescent, although, less anthropometric measurements were assessed (i.e., height, weight, waist circumference, triceps and subscapular skinfolds).

For the 2-km walk test the “time of performance” ranged between 11 to 23 minutes with a mean of 15.6 minutes for males and 16.7 minutes for females; similar results were found in previous studies [36, 37] in adults. In the 20-m shuttle run test, a range of 1.5 to 13 minutes with a mean of 6.3 minutes for males and 5.7 minutes for females. No studies have been found that record the “time of performance” in the adult population, but a previous study in preschoolers showed a range of 5.3 to 10 minutes [33–35] and a mean of 5.1 for children and adolescents [27]. It is important to note that the time spent in this study will depend on the cardiorespiratory fitness level of the participants, where sex and age are also determinants.

In general, the muscular fitness tests did not exceed 2 minutes in “time of performance”, including those with two attempts and 1-minute rest between attempts. Similar results have been found in two studies [27, 34], in which the “time of performance” for the handgrip strength and standing long jump (including 2 or 3 jumps and 1 minute rest between attempts) tests were ∼2 minutes in children and adolescents. Regarding the motor fitness tests, the 6-m gait speed and 2.45-m time up & go tests took less than 7 seconds (one attempt per test), the 4x10-m shuttle run test (including two attempts and 1-minute rest between attempts) took 1.2 to 1.3 minutes, and the single-leg stand test (30 seconds per leg and 1-minute rest between eyes open and eyes closed modality) took 3.0 to 3.6 minutes. Two studies that analysed the “time of performance” of the 4x10-m shuttle run test in preschoolers found a range from 1.3 to 1.4 minutes [34], and from 1.3 to 1.4 [35] (including two attempts and 1-minute rest between attempts). No other studies were found that examined this test in adults. Concerning the single-leg stand test, only one study [22] measured the “time of performance” in adults, using a 5-point scale. It was concluded that this test showed an excellent “time of performance” (i.e., range from 4.3 to 4.7).

### Safety

Referred to our safety assessments, >99% of the items presented a successful answer. No participant reported any “instrument allergy”, which is in line with a previous study in children and adolescents [27]. Regarding to “adverse events” item, there were no falls, and no participants suffered any injuries during the assessments; however, only one participant (0.3%) felt sick at the end of the 20-m shuttle run test, who recovered without complications after resting and hydrating. Only one study [39] in adults analysed “adverse events” item in the 6-min walk test and the results were similar to our study. Previous studies in adults observed a 0.5% [22] and <30% [37] of adverse events assessing the 2- km walk test; both expressed that the discomfort was focused on the lower limb musculature. Also, one study [35] reported no “adverse events”, and other study [27] showed than one participant reported muscle cramps during the assessment of the 20-m shuttle run test in children and adolescents. Regarding muscular fitness tests, 4 studies have included this item in the assessment of handgrip strength test. Three showed no adverse events in children [30, 35] and adults [22], and only one study [27] reported that one participant had “pain in the hand or forearm” during the assessment in children. Moreover, no adverse events were observed in the assessment of the standing long jump test in children [32, 35]. Finally, one study [22] in adults reported that three participants felt dizzy during the modified version of single-leg stand test (eyes-opened, 1 minute by legs) and one study [35] reported no adverse events in the 4x10-m shuttle run test in children. As we mentioned above, no “adverse events” occurred in our study, only the case of one participant who felt sick at the end of the 20-m shuttle run test but recovered without complications after resting and hydrating. In general, it may be noted that the incidence of adverse events in the field-based physical fitness tests is low and mostly related to strain of the predominant musculature involved in the performance of the test, which is something normally observed in physical exercise. This allows us to conclude that the application of these field-based physical fitness tests is safe.

Regarding to “HR” item, the most relevant aspect of monitoring it is the opportunity to assess the intensity of exercise, which is closely related to VO_2_ in submaximal (6-min walk and 2-km walk) and maximal (20-m shuttle run) field-based fitness tests, which allows to know accurately the intensity of these [40]. The “85%HRmax” is considered a rough estimate of the anaerobic threshold [41]. We observed that more participants reached “85% HRmax” as the test duration (i.e., the 6-min walk vs. the 2-km walk tests) and test intensity (i.e., 6-min walk and 2-km walk tests vs. the 20-m shuttle run test) increased. Similarly, with increasing age, more participants reached the “85%HRmax”, however, the level of physical activity of the participants did not influence reaching this threshold. This item is also used by Suni et al., [22] to calculate cardiovascular exertion in cardiorespiratory fitness test (i.e., the 2-km walk) as well as in muscular and motor fitness tests in adults (i.e., 37-57 years). Regarding the 2-km walk test, they reported that 43% of males and 37% of females reached or exceed their “85% HRmax”. In our study, the 32% reached their “85%HRmax”, being all the participants from the older groups (i.e., 35-49 and 50-64 years). This could mean that younger participants from our study (i.e., 18-34 years) would have better adaptations to the cardiovascular effort of this submaximal test. Only one study [36] reported the “HR” at the end of the 20-m shuttle run test in adolescents (177.7 (29.2) bpm), however, the “85% HRmax” of the participants was not reported. “HR” assessment has shown to be a useful measure of the cardiovascular stress level of participants [40]. For safety reasons, “HR” monitoring could be included in endurance strength tests like 30-s sit-to-stand and front plank tests.

Concerning to the item “RPE”, the results on perceived exertion in the muscular fitness tests (i.e., 30-s sit-to-stand and front plank tests) showed that the most demanding test for the participants was the front plank test, where more than 60% perceived their general exertion as “hard”. No other studies that registered “RPE” item in muscular fitness tests were found. We did not include “RPE” registering in the cardiorespiratory and motor fitness tests, however, others studies in adults analysed this item in cardiorespiratory fitness tests and showed “moderate” effort in the 2-km walk test [36, 37] and “very hard” effort in the 20-m shuttle run test [42]. We believe that the “RPE” can be also assessed in cardiorespiratory fitness tests, but it is essential to include an objective variable such as the “HR”.

In relation to “tibial pain” item, it was only registered after finishing the 2-km walk test, where the 32% of the total sample felt some degree of pain but any participant interrupted the test because of pain. There are no other safety studies that analyse this item. It is relevant to include the assessment of “tibial pain” as a method for quantifying muscle pain at the end of field-based walking tests (i.e., the 6-min walk and 2-km walk tests) in order to analyse the physiological response of the musculoskeletal system to the intensity and duration of walking tests. During the walking a higher pressure is generated on the tibialis anterior musculature, which is responsible for controlling the descent of the foot to floor contact and is involved in the dorsiflexion of the foot in the swing phase, so the higher intensity of walking may lead to the appearance of muscle pain [43, 44]. This assessment could be a good complement to the “DOMS” perceived by the participants.

We consider that it is relevant to include the “HR” and “RPE” items in cardiorespiratory and endurance strength fitness tests (i.e., 30-s sit-to-stand and front plank tests); and “tibial pain” in walking/running cardiorespiratory fitness tests, as they have been less analysed in previous studies, but are highly related to the control of the muscular and cardiovascular exertion of the participants and helps to quantify the exercise intensities during the performance of the field-based physical fitness tests [40, 45]. All this would provide further advances in the study of safety in health-related field-based physical fitness tests.

Referred to the “DOMS” item, in our study the 43.6% of the total sample (44.1% active) showed some degree of “DOMS”, and only the 2.9% reported a “severe” degree. From this sample, the 89.4% felt pain in lower extremities and additionally, the 95.3% of participants reported that cardiorespiratory fitness tests could be the cause of their “DOMS”. Only one study [22] assessed “DOMS” in adults indicating that 60% of males and 78% of females experienced some degree of “DOMS”, but only the 5% of males and 10% of females reported “severe” degree. Although this study also included the 2-km walk as a cardiorespiratory fitness test, 83% of participants reported that their “DOMS” were caused by the one-leg squat test. Another study [27] reported that the 35.3% of children and adolescents experienced some degree of “DOMS”, where the 21.3% reported “severe” degree and the 83% reported that the “DOMS” was due to performing the 20-m shuttle run test, which is in line with our study. It is important to include “DOMS” as a tool to detect the degree of muscular overload (intensity and/or duration) resulting from the assessment of the field-based physical fitness tests and also, for identify the muscular fitness of individuals in order to establish more personalized training loads related to gender, age and level of physical activity [46]. This tool, in addition to those mentioned above (HR, RPE and tibial pain), helps us to understand more precisely the physical demands of the most frequently used health-related field-based physical fitness tests in the adult population.

## Strength and limitations

This is the first study based on scientific evidence to determine the items to assess the degree of feasibility and safety of the most common health-related field-based physical fitness tests used in adults, which was conducted in a large and quite homogeneous sample in terms of sex, age and physical activity level. The main limitation in our study was not including the “RPE” registration in all the field-based physical fitness tests. This scale has shown to be a simple measure that can be used as a complement to “HR” monitoring for report the degree of exertion of the participants, particularly in the assessment of cardiorespiratory fitness tests.

## Conclusions

The most frequently used health-related field-based physical fitness tests are feasible and safe in adult population homogeneously distributed by sex, age, and physical activity level. The findings of the present study allow to determine what field-based physical fitness tests are feasible and safe in adults, necessary preconditions to propose a health-related field-based physical fitness test battery.

## Key Points

- This is the first study to determine that the most frequently used health-related field-based physical fitness tests are feasible and safe (items selected based on the science evidence) in adult population, homogeneously distributed by sex, age and physical activity level.
- We propose which items should be used to assess feasibility: “appropriate facilities”, “appropriate equipment”, “appropriate sport clothes”, “test’s instruction understood”, “easy to administer”, “participants evaluated/not evaluated”, ratio participants/evaluators”, “time of preparation”, and “time of performance”.
- The items to be used to assess safety should be: “instrument allergy”, “adverse events” (i.e., falls, injury and sick feeling), “heart rate”, “85% heart rate max”, “rate of perceived exertion”, “tibial pain”, and “delayed-onset muscle soreness”.
- We consider that it is relevant to include the “heart rate” and “rate of perceived exertion” items in cardiorespiratory and endurance strength fitness tests (i.e., 30- s sit-to-stand and front plank tests); and “tibial pain” in walking/running cardiorespiratory fitness tests.

## Data Availability

All relevant data are included in the article and/or its supplementary information files.

## Acknowledgments

We thank all participants and evaluators who took part in the ADULT-FIT study.

## Statements and Declarations

### Author contributions

CCL, JCP and MCG contributed to the study in terms of conception, design and critical review. CCL led the writing of the study and NMJ contributed to the methodology. All authors discussed the results and contributed to the final manuscript and agreed on the order of presentation of the authors. All authors have read and approved the final manuscript.

## Availability of data and material

The authors declare that all relevant data are included in the article and/or its supplementary information files.

## Informed consent

All participants provide voluntarily written informed consent to be part of the present study. The study was approved by the Committee for Research of Cadiz, Spain.

