## Supplementary Material 1 for "Feasibility and Safety in health-related field-based physical fitness tests in adult population: The ADULT-FIT project"

[c]PA-HELP "Physical Activity for Health Promotion" research group. University of Granada, Granada, Spain.

***Corresponding author:** Carolina Cruz-León, GALENO research group, Department of Physical Education, School of Education, University of Cádiz, Puerto Real, Spain. Avenida República Saharaui s/n, 11519, Puerto Real (Cádiz), Spain. Telephone number: +34 956 016 253

**Supplementary Material 1**

As mentioned in the manuscript, the tests were carried out in three non-consecutive days, with at least 72 hours apart, in an indoor/outdoor facility under appropriate conditions. The tests assessments were distributed in the following order: day 1) Body composition measurements, the 4x10-m shuttle run, handgrip strength, and 2-km walk tests; day 2) the single-leg stand, standing long jump, front plank, and 6-min walk tests; day 3) the 2.45-m time up & go, 6-m gait speed, 30-s sit-to-stand, and 20-m shuttle run tests.

**Body composition**

The body composition measurements included weight, height, neck, waist and hip circumferences, subscapular and triceps skinfolds, following the protocol described by the International Society for the Advancement of Kynanthropometry [1]. The protocols established by the Centers for Disease Control and Prevention were followed to assess neck circumference [2]. All measurements were taken by the same trained evaluator (to avoid intra-evaluator variability) twice and the mean between measurements was calculated. The evaluator was of the same sex as the participant.

**Motor fitness**

*6-m gait speed test*

The participants had to cover a distance of 6 metres walking as fast as possible without stopping. This test has a total distance of 10 metres. In the first 2 metres the participant will accelerate his/her walking to the maximum, and for the next 6 metres, the participant will walk as fast as possible (recorded distance). Afterwards, the participant will have two metres to decelerate. The test was evaluated and supervised by two evaluators, one of them located at the starting line of the course (point 1) and the second one, lateral to the 6 metres line (point 2), measuring the time of execution [3]. One attempt was performed.

*2.45-m time up & go test*

The test assesses mobility skills, gait speed and functional ability. Participants started the test sitting on a chair with both arms crossed over the chest. The participant had to get up from the chair, walk 2.45-m (8 feet) as fast as possible, turn, and return to seated position. The test was evaluated and supervised by two evaluators; one of them gave the start signal and controlled the total time required for the participant to complete the out and back. The second one held the chair of the participant at the time of return to prevent possible incidents. One attempt was performed.

*4x10-m shuttle run test*

The aim of this test was to assess the speed of movement, coordination and agility. It is a run and turn test at maximum speed (4x10-m) between two parallel lines on the floor 10 metres apart. On the start line there was a sponge (B) and on the opposite line there were two sponges (A, C). At the signal, the participant ran without a sponge as fast as possible to the other line crossing the line with both feet, picking up the sponge A and returning to the starting line. Quickly changing sponge A for sponge B and running back as fast as possible to the opposite line and changing sponge B for sponge C, returning to the starting line and finishing the test. The test was evaluated and supervised by one evaluator located at the start/end line, who recorded the time spent by the participant [4]. Participants had to perform two non-consecutive attempts and the best attempt was recorded.

*Single-leg stand test (eyes open and closed)*

The aim of the test was to assess the postural balance over a reduced support area. The test was performed by supporting on one foot and raising the opposite leg in a tandem position with their arms at their sides parallel to their trunk and with their eyes open during 30 seconds; and subsequently with the other leg. Participants also performed the test with eyes closed for 30 seconds per leg. The test was evaluated and supervised by two evaluators. Both recorded every time the participant lost balance and put his/her foot on the floor, or if the participant performed any support on the leg that was steady. When the test was performed with eyes closed, one of the evaluators recorded each time the participant opened his eyes and the second recorded the losses in balance. It was the duty of the evaluators (when required), to ask participants to maintain the balance position by not supporting the tandem leg on the floor or on the balance leg [5]. The test was performed once per leg, first with eyes open and then with eyes closed. Two non-consecutive attempts of each round were evaluated.

**Muscular fitness**

*Handgrip strength test*

The aim of the test was to assess the maximal isometric strength according to the protocol described by Ruiz et al., [6], where in the case of women, the grip had to be adjusted according to the size of the hand. Participants had to squeeze the dynamometer continuously and gradually for at least 3 seconds. The elbow should be in full extension and slightly away from the body to avoid contact of the dynamometer with any other part of the body. The test was evaluated and supervised by one evaluator. Two non-consecutive attempts were made with each hand and the best result at each grip span was retained.

*Standing long jump test*

The aim of the test is to assess the lower body explosive strength through an energetic jump forward in an attempt to reach the greatest distance possible without losing balance. Participants had to jump with both feet from a marked line to the front, trying to reach as far as possible with both feet together. The test was evaluated and supervised by one evaluator. Two non-consecutive attempts were performed and the best attempt was recorded [7].

*30-s sit-to-stand test*

The aim of this test was to assess the lower body endurance strength to sit down and stand up for 30 seconds as fast as possible while maintaining a correct execution throughout the test. Participants were seated in the centre of a chair, back straight, with their arms crossed on their chests, legs approximately shoulder-width apart, feet placed on the floor at a 90-degree angle to the knees and slightly in front of the chair legs to avoid possible impact when rising up. The participants had to stand up and sit down as many times as possible for 30 seconds. The test was evaluated and supervised by two evaluators, one of them holding the chair and registering the rate of perceived exertion (RPE) using a Borg scale (1-10 points) [8] at the end of the test, and the second one checking the time and number of repetitions performed by the participant [9]. One attempt was performed.

*Front plank test*

The aim of the test was to assess the trunk endurance strength by performing a frontal plank for as long as possible. The starting position of the test was with the knees, toes and elbows resting on the floor, the palms of the hands in contact with the floor, maintaining a neutral position of the cervical curvature and forming a straight line between the lateral area of the acromion, greater trochanter and malleolus. The forearm was at a 90º angle to the upper arm and the hands were placed shoulder width apart. Prior to the test, an elastic cord was attached horizontally to a pair of vertical sticks, which was moved to a height that was at the same level as the participant's iliac crest, and then adjusted 3 centimetres below. This adjustment served as a reference for objective control of hip displacement during the test. Each time the hip touches the reference line, a verbal warning from the assessors would be given so that the participant recovers the correct position. The test would be finished when the hip was not maintained at the required level after receiving two consecutive warnings or when the participant reached the maximum effort. At the end of the test, total time and the RPE was registered. The test was evaluated and supervised by two evaluators, who were positioned one on each side of the participant to ensure that the participant maintained the correct position and tracked the time performed with a hand-held stopwatch (Casio HS-EV-1RET Digital, Casio, Tokyo, Japan) [10]. One attempt was performed.

**Cardiorespiratory fitness**

The cardiorespiratory field-based physical fitness tests assessed were: i) 6-min walk [11], ii) 2-km walk [12] and iii) 20-m shuttle run [13]. All the tests were performed in groups of 10 participants approximately.

*6-min walk test*

The aim of the 6-min walk test was to walk and reach the longest distance possible in 6 minutes without running and keeping a steady pace. The participants started the test simultaneously, walking in a space of 20 x 30 metres. Two evaluators monitored the time and subsequently recorded the total distance covered, and the heart rate (HR) of the participants at the end of the test. One attempt was performed.

*2-km walk test*

For the 2-km walk, the aim of the test was to complete the 2 kilometres distance walking in the shortest time possible without running and keeping a steady pace. The participants started the test simultaneously, walking in a space of 20 x 30 metres. Two evaluators recorded the number of laps (participants had to complete 20 laps) during the 2-km walk test, total time, HR and tibial pain through a numeric rating scale (0-10 points) at the end of the test. One attempt was performed.

*20-m shuttle run test*

For the 20-m shuttle run test, participants had to run between 2 lines 20 metres apart following the rhythm of an audible signal. The initial speed of the signal was 8.5 km/h, which was increased by 0.5 km/h/min (1 minute equaled 1 stage), so the participants had to rise their running pace to reach the corresponding line. The test was finished when participants reached their maximum effort and abandoned the test or failed to reach the end lines at the same time as the audio signals on two consecutive occasions. Two evaluators registered the stages performed by each participant and the HR at the end of the test. One attempt was performed.

The HR was recorded using the activity bracelet Xiaomi Mi Band 4 (Xiaomi Inc, Beijing, China) validated in adults [14].
